# COMPARISON OF sPLA2-IIA PERFORMANCE WITH HIGH-SENSITIVE CRP, NEUTROPHIL PERCENTAGE, PCT AND LACTATE TO IDENTIFY BACTERIAL INFECTION: A PROSPECTIVE STUDY

**DOI:** 10.1101/2021.03.02.21252745

**Authors:** Toh Leong Tan, Christabel Wan-li Kang, Kai Shen Ooi, Swee Thian Tan, Nurul Saadah Ahmad, Dian Nasriana Nasuruddin, Azlin Ithnin, Khaizurin Tajul Arifin, Yook Heng Lee, Nurul Izzaty Hassan, Kok Beng Gan, Hui-min Neoh

**Author notes:** Universiti Kebangsaan Malaysia Medical Centre, 56000 Cheras, Kuala Lumpur, Malaysia (Toh Leong Tan, Kai Shen Ooi, Swee Thian Tan, Nurul Saadah Ahmad, Dian Nasriana Nasuruddin, Azlin Ithnin, Khaizurin Tajul Arifin, Hui-min Neoh). Tengku Ampuan Rahimah Hospital, Ministry of Health, 41200 Klang, Selangor, Malaysia (Christabel Wan-li Kang). Universiti Kebangsaan Malaysia, 43600 Bangi, Selangor, Malaysia (Yook Heng Lee, Nurul Izzaty Hassan, Kok Beng Gan).

## Abstract

Early bacterial infection (BI) identification in resource-limiting Emergency Departments (ED) is challenging especially in low- and middle-income counties (LMIC). Misdiagnosis predisposes to antibiotic overuse and propagates antimicrobial resistance. This study evaluates new emerging biomarkers, secretory phospholipase A2 group IIA (sPLA2-IIA), and compares with other biomarkers on their performance characteristic of BI detection in Malaysia, an LMIC. A prospective cohort study was conducted involving 151 consecutive patients admitted to the ED. A single measurement was taken upon patient arrival in ED and was analysed for serum levels of sPLA2-IIA, high-sensitive C-reactive protein (CRP), procalcitonin (PCT), neutrophil percentage (N%), and lactate. All biomarkers’ performance was compared for the outcomes using area under the receiver operating characteristic curve (AUROC), sensitivity, and specificity. The performance of sPLA2-IIA (AUROC 0.93 [95% CI: 0.89-0.97]; Sn 80% [95% CI: 72-87] Sp 94% [95% CI: 81-89]) was the highest among all. It was comparable with high-sensitive CRP (AUROC 0.93 [95% CI: 0.88-0.97]; Sn 75% [95% CI: 66-83]; Sp 91 [95% CI: 77-98]) but had a higher Sn and Sp. The sPLA2-IIA was also found superior to N%, PCT, and lactate. This finding suggested sPLA2-IIA was recommended biomarkers for BI detection in LMIC.

## INTRODUCTION

The Global Burden of Disease Study reported that infection caused more than 10 million lives lost per year.^1^ Particularly, pneumonia is one of the principal causes of morbidity and mortality in adults and children, with its epidemiology varies markedly by region. Various studies show a marked difference in mortality rate from high-income to low- and middle-income countries (LMIC), with the highest death, observed in Sub-Saharan Africa and Southeast-Asia (SEA).^2-4^ In Malaysia, an LMIC in SEA, pneumonia remains amongst the leading cause of death right after ischemic heart disease since 2014 across all age groups, gender, ethnicities, and stratum.^5^

Infection, not limited to pneumonia, can happen elsewhere caused by a diversity of microbial pathogens. Hence, identification of the aetiological agent is the key in determining patient recovery. It is clinically essential to differentiate bacterial infection (BI) and non-bacterial infection (NBI) as the treatment protocol differs significantly. Nevertheless, considering the similar clinical presentation between bacterial and viral infections, it could be challenging in distinguishing the two based on both history taking and examinations.^6^

Theoretically, by isolating out the causative agents, blood culture is always recognised as the gold standard to diagnose BI. The most drawback is the significant turnaround time of approximately 24-48 hours to isolate the causative agents. This time-intensive limitation had become the disadvantages for blood culture and rendering it impracticable at ED for its triage responsibility.^7,8^ Hence, the delay in diagnosis would render the initiation of empirical antibiotics. Saleh, et al. illustrated that up to 30% of the clinicians proceed to continue the antibiotic, although the results were yet to confirm bacterial infection.^7^ Frequent abuse and misuse of the antibiotic attribute to antimicrobial resistance becoming one of the biggest threats to global public health. Moreover, individual patient health is at stake with prolonged hospitalisation, radical treatment, and soaring healthcare expenses.^6^

Therefore, there is an ongoing effort to pursue a simple and accurate diagnostic tool. Biomarkers appear as a promising point-of-care test in the triage system. Culture and sensitivity would play a supportive role to further assist in proper clinical judgement. Among the biomarkers, CRP and PCT are the most extensively studied. However, most of them being studied in high-income countries with limited data generated from LMIC.^9^ Nevertheless, the performances were somewhat inconsistent and fluctuated.^10,11^ For the new emerging biomarkers, secretory phospholipase A2 group IIA (sPLA2-IIA) was hypothesised able to distinguish BI, but few studies were available, hence requiring further validation.^12,13^ This prospective study aimed to investigate the performance of new emerging biomarker sPLA2-IIA with other biomarkers inclusive of high-sensitive CRP, PCT, N%, and lactate in their diagnostic value to identify BI from NBI.

## RESULTS

### CHARACTERISTIC OF STUDY COHORT

From March 2015 – October 2017, a total of 154 patients admitted to the Emergency Department were who fulfilled the inclusion criteria, of which 151 consecutive patients were eligible and selected for this study after exclusion of three missing blood samples (**Figure 1**). The demographic data of the patient characteristics were shown in **Table 1**, and causative pathogen data was presented in **Table 2**. The mean age was around 57.8 years old with equal gender distribution. Among the 151 patients, 115 were diagnosed to have BI. All biomarker results were compared at a cut-off level determined via AUROC analysis.

**Table 1:** Sociodemographic data of patients recruited for the study.

| **Demography ( n=151 )** |  |  |
| --- | --- | --- |
| **Age (years; mean [SD])** | 57.8 | (19.3%) |
| **Gender, No. (%)** |  |  |
| Male | 80 | (53.0%) |
| Female | 71 | (47.0%) |
| **Race, No. (%)** |  |  |
| Malay | 67 | (44.0%) |
| Chinese | 63 | (42.0%) |
| Indian | 9 | (6.0%) |
| Others | 12 | (8.0%) |
| **Bacterial infection** | 116 | (77.0%) |
| **Non-bacterial infection** | 35 | (23.0%) |
| **Source of infection, No. (%)** |  |  |
| Respiratory | 48 | (31.8%) |
| Urinary | 27 | (17.9%) |
| Musculoskeletal | 22 | (14.6%) |
| Gastrointestinal | 14 | (9.3%) |
| Skin | 13 | (8.6%) |
| Cardiac | 10 | (6.6%) |
| Dengue | 7 | (4.6%) |
| Central nervous system | 3 | (2.0%) |
| Blood/Catheter related | 3 | (2.0%) |
| Obstetrics & Gynaecology | 2 | (1.3%) |
| Thyroid | 1 | (0.7%) |
| Eye | 1 | (0.7%) |

**Table 2:** Causative microbial detected via cultivation.

| Cultured organisms | Frequency | Percentage |
| --- | --- | --- |
| *Escherichia coli* | 15 | (9.9%) |
| *Dengue virus* | 7 | (4.6%) |
| *Staphylococcus aureus* | 6 | (4.0%) |
| *Proteus* spp. | 4 | (2.6%) |
| *Pseudomonas aeruginosa* | 4 | (2.6%) |
| *Candida* spp. | 3 | (2.0%) |
| *Escherichia coli* ESBL<sup>a</sup> | 3 | (2.0%) |
| *Klebsiella* spp. ESBL<sup>a</sup> | 3 | (2.0%) |
| *Streptococcus* Beta-Hemolytic Group B | 3 | (2.0%) |
| *Bacteroides* spp. | 2 | (1.3%) |
| *Klebsiella* spp. | 2 | (1.3%) |
| Methicillin-resistant *Staphylococcus aureus* | 2 | (1.3%) |
| *Morganella morganii* | 2 | (1.3%) |
| *Mycoplasma tuberculosis* | 2 | (1.3%) |
| *Streptococcus* Beta-Hemolytic Group G | 2 | (1.3%) |
| *Streptococcus viridans* | 2 | (1.3%) |
| *Burkholderia pseudomallei* | 1 | (0.7%) |
| *Chlamydia Pneumoniae* | 1 | (0.7%) |
| *Enterobacter* spp. | 1 | (0.7%) |
| *Enterobacter* spp. ESBL<sup>a</sup> | 1 | (0.7%) |
| *Enterobacter cloacae* | 1 | (0.7%) |
| *Klebsiella pneumoniae* | 1 | (0.7%) |
| *Klebsiella pneumoniae* ESBL<sup>a</sup> | 1 | (0.7%) |
| *Salmonella* spp. | 1 | (0.7%) |
| *Scytalidium* spp. | 1 | (0.7%) |
| *Staphylococcus coagulase negative* | 1 | (0.7%) |
| *Streptococcus pneumoniae* | 1 | (0.7%) |

<sup>a</sup>Extended-spectrum beta-lactamases

**Figure 1:**
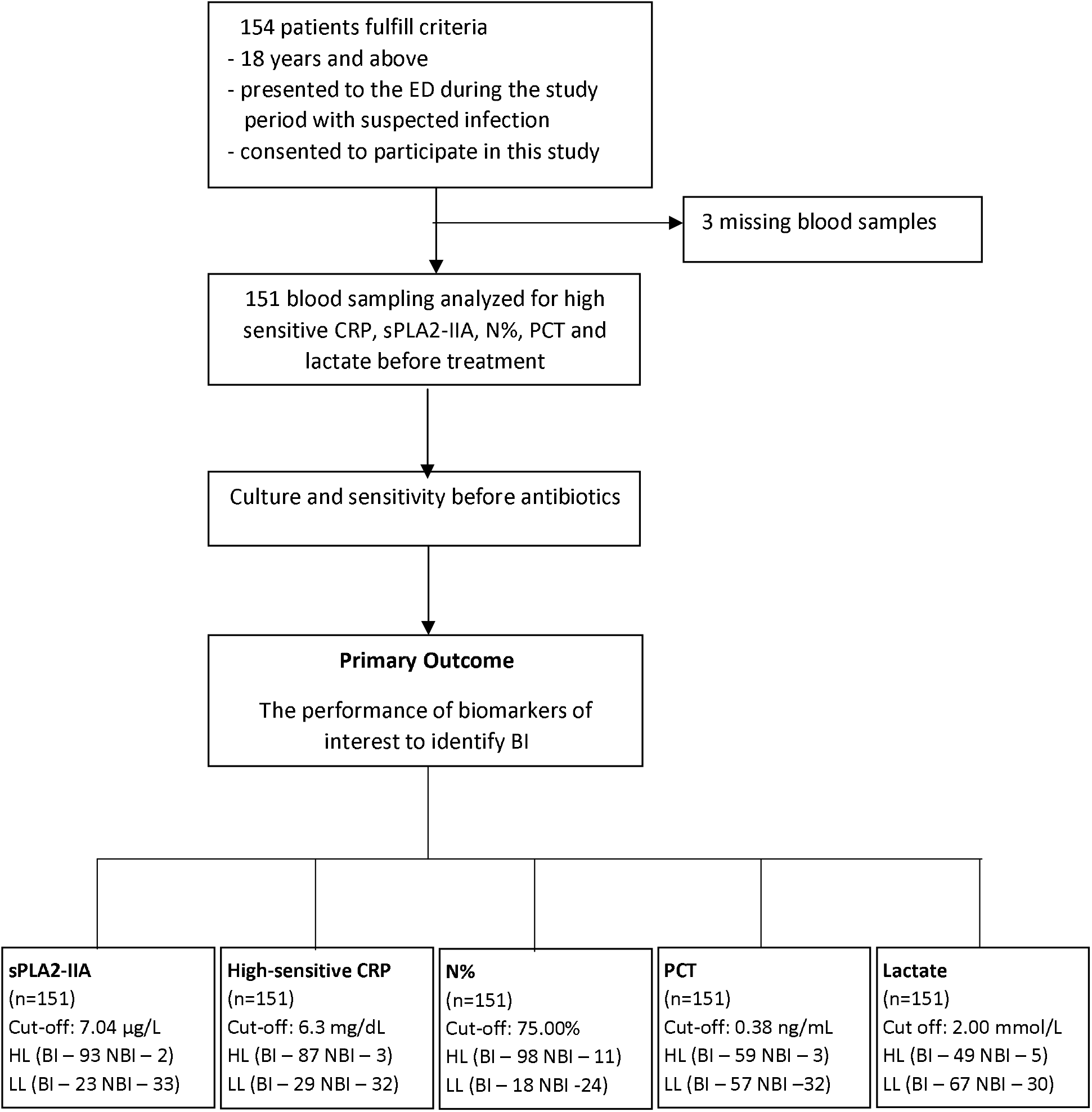
Study on-site recruitment and workflow (May 2015-October 2017). High-sensitive CRP = High-Sensitive C-Reactive Protein; sPLA2-IIA = Secretory Phospholipase 2-IIA; N%, = Neutrophil Percentage; PCT = Procalcitonin; HIV = Human Immunodeficiency Virus; ESRF = End-Stage Renal Failure; BI = Bacterial Infection; NBI = Non-Bacterial Infection; HL = High Level*; LL = Low Level^†^ * High level refers to biomarker’s level that equal to or higher than the cut-off point † Low level refers to biomarker’s level that equal to or lower than the cut-off point

### DETECTION OF BACTERIAL INFECTION

According to BI and NBI groups, the median and interquartile range for all biomarker levels were summarised in **Table 3** and **Figure 2**. All biomarker median levels were significantly higher in the BI group than the NBI group (*P*<0.001). The AUROC values, Sn, Sp, PPV, NPV, accuracy, and κ for all biomarkers for detecting sepsis and BI were shown in **Table 4** and **Figure 3. Table 3** showed that high-sensitive CRP and sPLA2-IIA were found to have the highest AUROC values, both 0.93 (95% CI, 0.88-0.97). **Figure 3** presents the AUROC for all biomarkers in differentiating BI. Interestingly, sPLA2-IIA was found to have the best Sp (94%, 95% CI, 0.81-0.99) with a cut-off point of 7.04 μg/l. N% had the highest Sn (84%, 95% CI, 0.77-0.91) among all the biomarkers, with a cut-off of 75%. High-sensitive CRP, PCT, and sPLA2-IIA had higher cut-off points in BI detection than in sepsis detection. In further analyses of AUROC among biomarkers, high-sensitive CRP, sPLA2-IIA, and N% predictability of BI were found equally good (*P*>0.05). These three biomarkers also had significantly higher AUROC than PCT and lactate (*P*<0.05) **(Table 5)**

**Table 3:**
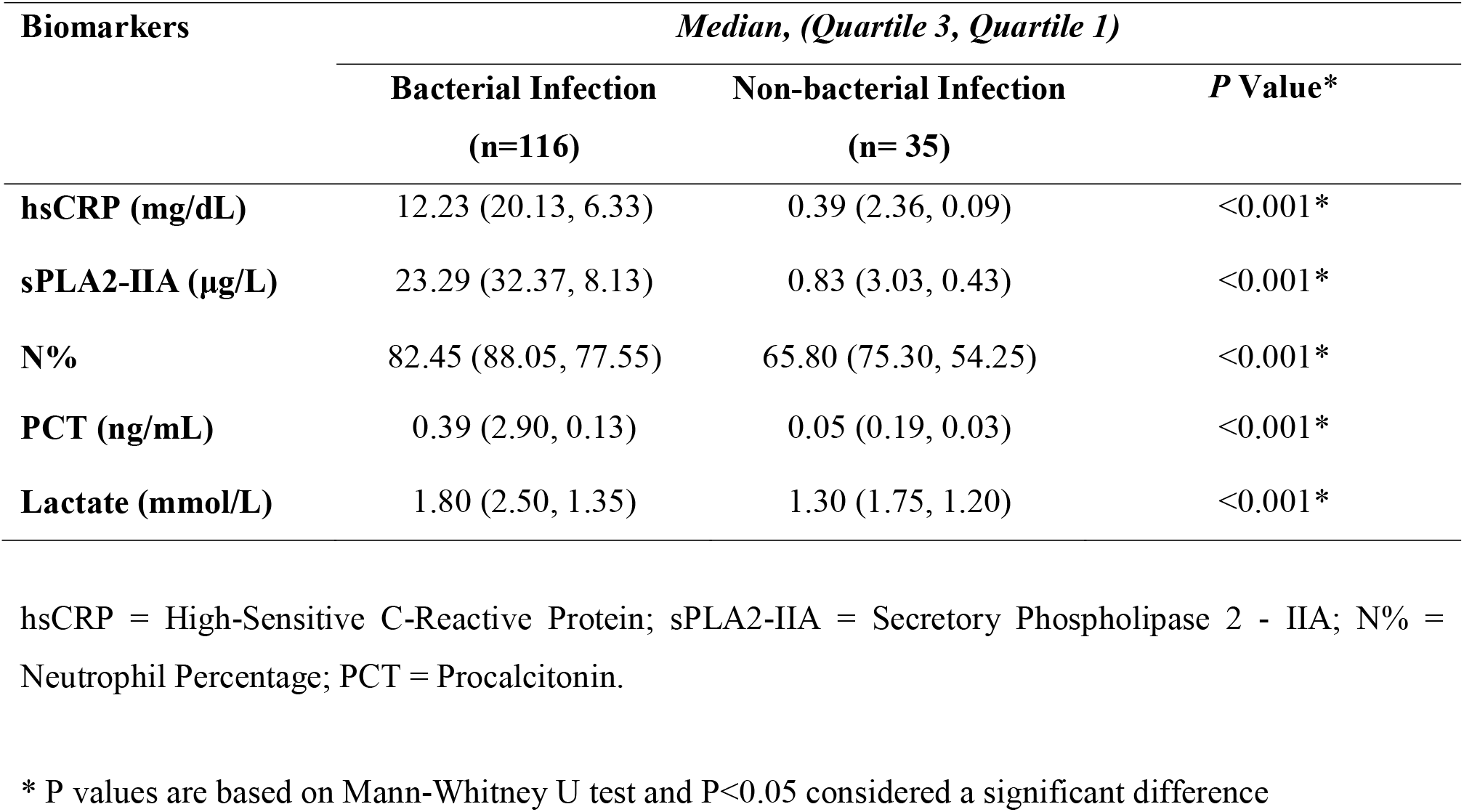
Comparison of high-sensitive CRP, sPLA2-IIA, N%, PCT and lactate levels in Bacterial Infection, Non-bacterial Infection groups of this study.

**Table 4:** The ability of five tested biomarkers to differentiate bacterial infection in ED.

| **Biomarker** | **AUROC\*†**<br>**(95%CI)** | **Cut-off**<br>**point** | **Sn (%)**<br>**(95%CI)** | **Sp (%)**<br>**(95%CI)** | **PPV (%)**<br>**(95%CI)** | **NPV (%)**<br>**(95%CI)** | **Kappa (κ)** | **P Value§** | **Accuracy**<br>**(95%CI)** |
| --- | --- | --- | --- | --- | --- | --- | --- | --- | --- |
| **Bacterial (n=116) versus non-bacterial infection (n=35)** |  |  |  |  |  |  |  |  |  |
| **hsCRP (mg/dL)** | 0.93 (0.88-0.97) | 6.33 | 75 (66-83) | 91 (77-98) | 97 (91-99) | 52 (44-61) | 0.53 | <0.001 | 79 (71-85) |
| **sPLA2-IIA (µg/L)** | 0.93 (0.89-0.97) | 7.04 | 80 (72-87) | 94 (81-99) | 98 (92-99) | 59 (50-68) | 0.62 | <0.001 | 83 (77-89) |
| **N%** | 0.85 (0.76-0.93) | 75.00 | 84 (77-91) | 69 (51-83) | 90 (84-94) | 57 (45-68) | 0.50 | 0.001 | 81 (74-87) |
| **PCT (ng/mL)** | 0.81 (0.73-0.89) | 0.38 | 51 (41-60) | 91 (77-98) | 95 (87-98) | 36 (31-41) | 0.28 | <0.001 | 60 (52-68) |
| **Lactate (mmol/L)** | 0.68 (0.59-0.77) | 2.00 | 42 (33-52) | 86 (70-95) | 91 (81-96) | 31 (27-36) | 0.17 | 0.002 | 52 (44-61) |

AUROC = Area Under Receiver Operating Characteristic Curve; Sn = Sensitivity; Sp = Specificity; PPV= Positive Predictive Value; NPV = Negative Predictive Value; hsCRP = High-Sensitive C-Reactive Protein; sPLA2-IIA= Secretory Phospholipase 2 - IIA; N% = Neutrophil Percentage; PCT = Procalcitonin.
\*AUROC classification: 0.70-0.79=Fair, 0.80-0.89=Good, >0.90=Excellence.
† Patients recruited excluding missing data(biomarkers) patients; complete case analysis
§ P values are based on $\chi^2$ test, $P<0.05$ indicates a significant difference

**Table 5:**
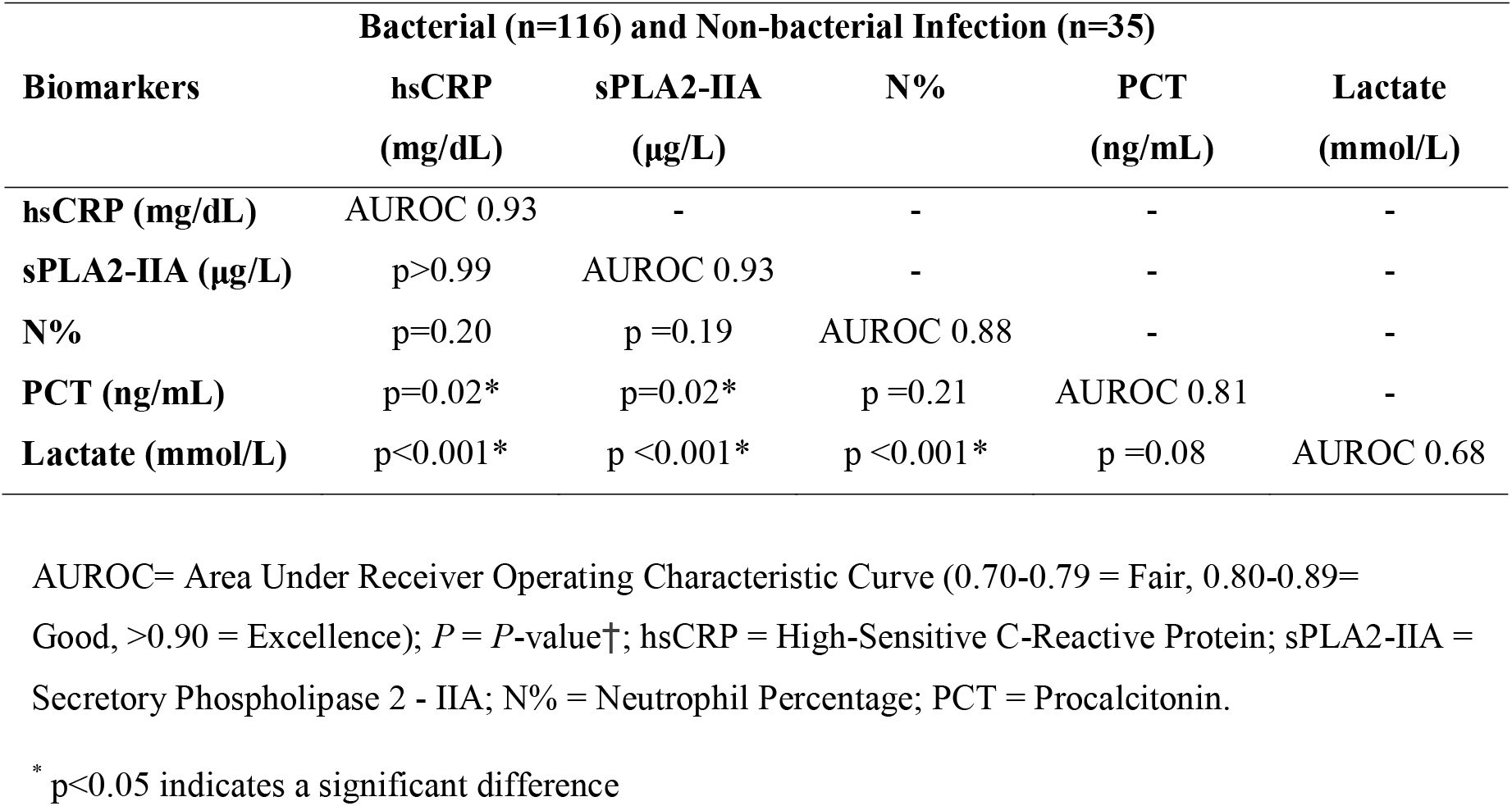
Comparison of The Performance of Biomarkers’ AUROC to Differentiate Bacterial Infection in ED.

**Figure 2:**
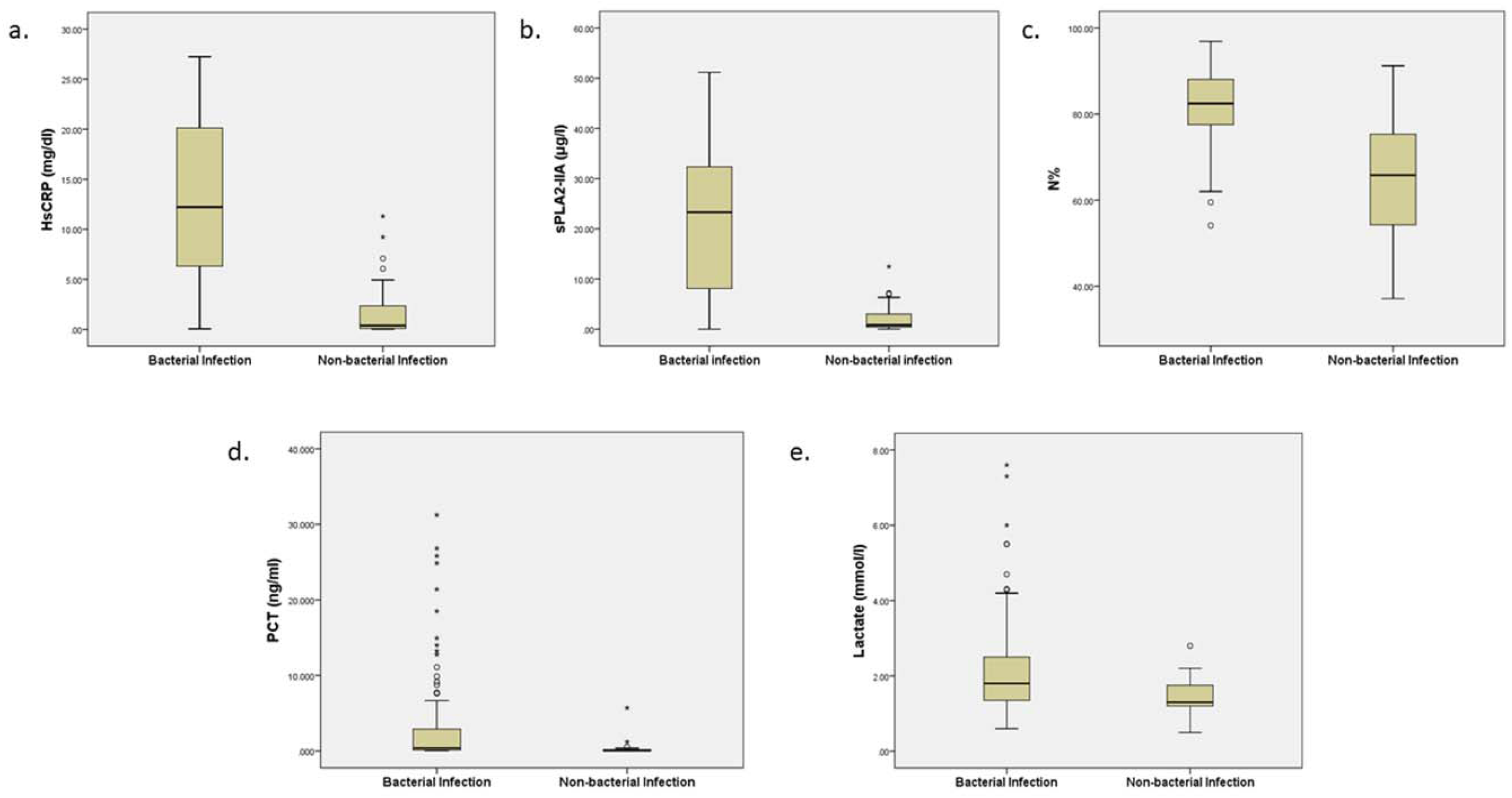
Box Plots for high-sensitive CRP, sPLA2-IIA, N%, PCT, and Lactate Levels in Detecting Bacterial Infection. hsCRP = High-Sensitive C-Reactive Protein; sPLA2-IIA = Secretory Phospholipase 2-IIA; N% = Neutrophil Percentage; PCT = Procalcitonin Boxes show the 25^th^-75^th^ centiles, while whiskers indicate the 10^th^ and 90^th^ centiles. Horizontal lines within the boxes indicate the median. Outliers are shown as circles and stars.

**Figure 3:**
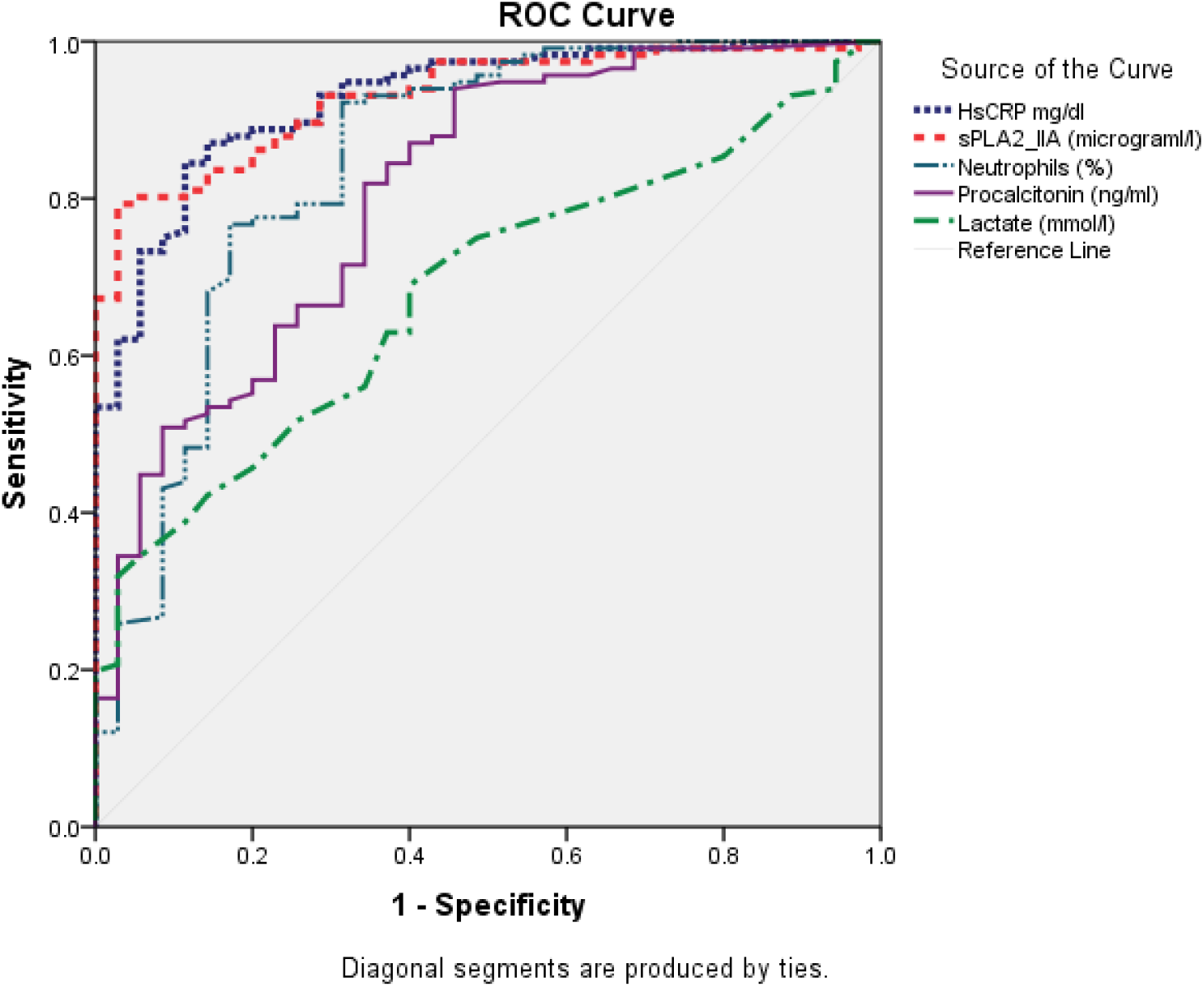
AUROCs of Five Tested Biomarkers in Differentiating Bacterial Infection in ED. This figure showed that all biomarkers able to predict sepsis and bacterial infection (BI). High-sensitive CRP, sPLA2-IIA, and N% have the highest AUROC among all for both sepsis and BI detection in ED. AUROC = Area Under Receiver Operating Characteristic Curve; hsCRP = High-Sensitive C-Reactive Protein; sPLA2-IIA = Secretory Phospholipase 2-IIA; N% = Neutrophil Percentage; PCT = Procalcitonin

## DISCUSSION

In our study, high-sensitive CRP and sPLA2-IIA had better AUROCs followed by N%, PCT, and lactate in discriminating sepsis and differentiating BI.

Despite limited literature available, the sPLA2-IIA performance in our study parallel with the findings from Rintala et al. (1993), showing that sPLA2-IIA was significantly higher in BI patients.^14^ The same study also highlighted a strong correlation of sPLA2-IIA level to high-sensitive CRP and PCT. In our current study, we found sPLA2-IIA has the equal performance to high-sensitive CRP and outweigh PCT. This also further justify with Tan et al. (2017). The primitive role of sPLA2-IIA coined as a bactericidal enzyme, catalysed bacterial membranes’ hydrolysis.^13^ This acute-phase protein engages the body host in response to inflammation and generates pro-inflammatory metabolites.^12^ Hence, its activity level is reliable to measure the degree of systemic inflammation in various bacteremic and non-bacteremic infections and differentiate between bacterial and viral infections.^12,15^

Several studies comparing CRP and PCT in the detection of BI showed that PCT was superior to CRP;^16-20^ however, our results showed otherwise. Postulation to this situation regards the high prevalence of co-morbidities in LMIC populations such as HIV, malaria, parasites, and malnutrition. The confounding chronic backgrounds influenced the molecules’ expression levels, thus skewing the assessed biomarkers’ levels.^21^ As a result of this, the authors highlighted the necessity to appraise biomarkers in LMIC populations for its intended application settings, considering there was a lack of reference tests for comparative analysis. Our CRP cut-off point falls within the cut-off range above 60-80 mg/dL that bacterial infection may be present.^8^ We hope the biomarkers value and their cut-off point in our study could serve as a reference and guidance for future result interpretation among the LMIC countries. Being the leading most extensive study on CRP and PCT’s efficacy for predicting bacterial infection in tropical, malarial endemic settings in the Southeast Asian context, Lubell et al. (2015, 2016) concluded that CRP outperforms PCT in its accuracy.^22,23^ Although the role of CRP was debatable as a biomarker of either inflammation or infection,^7,8^ our result affirmed the stand of Escadafal et al. (2020) whereby CRP levels correlated with the presence of bacterial infection and was consistent across various studies.^24^ Our study showed better performance of CRP than PCT which is comparable to other LMIC context studies in contrary to the observations in high-income countries.^10,23,25^

PCT played an essential role in antibiotic stewardship (AMS) and was recognised internationally for both its diagnostic and prognostic properties.^26,27^ The efficacy was validated in several trials showing a decreased antibiotic prescription rate and improvement in patients’ clinical outcomes. The Berlin 2018 expert consensus had developed a PCT-guided AMS concerning illness severity and likelihood for BI. Subsequently, algorithm adaptations were made to harmonies the PCT usage across the Asia-Pacific region given the differences in LMIC background. Nevertheless, the modified consensus also agreed that the PCT-guided AMS was not applicable in patients with suspected tropical disease.^27^ From the Berlin 2018 consensus, the PCT cut-off was fixed at 0.5 ng/mL at the intensive care unit (ICU) and 0.25 ng/mL non-ICU setting to predict the likelihood of BI.^26^ However, our study reported a higher PCT cut-off at 0.38 ng/mL as compared to the non-ICU group. As ED stands a special setting that harbours a mixture of critical and non-critical ill patients, and hence a higher PCT cut-off is found to be more practical for BI identification. Furthermore, the uprising performance in sPLA2-IIA compared to PCT in our study prompts the consideration of sPLA2-IIA to rise as a potential AMS biomarker. This current study encourages further exploration of sPLA2-IIA on AMS as an approach for more judicious antibiotic usage in the future time.

In previous literature, N% showed weak BI prediction in elderly patients. With an 80% cut-off point, low Sn and Sp were recorded at 35% and 74%, respectively.^28^ Comparatively, our study demonstrated a slightly lower N% cut-off (75%) with a higher Sn (84%) and Sp (69%). Although it is not the best biomarker, N% showed promising performance and outperformed PCT and lactate. Such attribution may relate to the neutrophils’ role as the predominant immune cell population migrated to the affected site regardless of the etiological agents at the early stage of infection. Mainly, the neutrophils were markedly increased during bacterial or fungal infections as compared to viral infections.^11^ Considering N% as an inexpensive and readily available biomarker, it promised to be an excellent parsimonious biomarker in detecting BI in LMICs. Therefore, a high N% should prompt a high BI suspicion and warrant further investigation in our settings.

In contrast, our study found that lactate was poor in differentiating BI, even though it has been used widely in clinical settings as a biomarker to discriminate sepsis from non-sepsis patients. Lactate reflects strained cellular metabolism as it is an end product in anaerobic metabolism from hypoxemia. A wide variety of conditions, such as trauma, endocrine emergency, acute cardiac events, and increased bacterial load, lead to lactate level elevation, impeding its sensitivity.

This study addressed several limitations that merit consideration. However, it was a single-centre ED-based study with all data samples collected solely from an academic medical center. Hence, selective bias may have occurred, and this study result may not apply to other hospitals’ ED settings. Apart from that, the study excluded the minorities with the inclusion age criterion restricted to those aged ≥ 18 years old. The paediatric population appeared to be an interesting group for further investigation as its application may possess considerable clinical potential and relevance. Undoubtedly, this will potentially support the use of biomarkers in all age groups to identify patients with BI. Lastly, the available clinical algorithm and lab test for microbiological diagnosis may be imperfect. The diagnostic performance may have negatively affected several cases whereby bacterial infection was misclassified as other pathogenic agents at some instances in which patients harbour bacterial infections without being microbiologically proven. Regardless, antibiotics would still be administered in these cases as part of a regular clinical routine. Our study’s strength is that the prospective study was carried out as proposed despite limited resources in our setting. Our study had achieved the proposed sample size, which ensured a high-power study.

## METHODS

### STUDY DESIGN, POPULATION AND SETTING

The research was approved by the Universiti Kebangsaan Malaysia (UKM) ethical review board (ethic code FF-2015-322) and carried out according to Good Clinical Practice guideline. This single-centre prospective cohort study was conducted over 30 months (May 2015-October 2017) in ED UKM Medical Centre, Malaysia. The ED UKM Medical Centre, with 72000 ED visits annually, served as an urban, academic teaching hospital with 1000-beds. The target population includes ≥ 18 years old who presented to the ED suspicious of infection throughout the study. Patients enrolled had given their written informed consent to participate in this study. The diagnosis of infection was determined by attending ED physicians based on the criteria stated in Horan et al. (2008).^29^ The patients were then be grouped into either BI or NBI. The bacterial infection is defined as a clinical bacterial infection or positive bacterial cultures (sputum, body fluids, blood, et cetera). However, the exclusion criteria were immunosuppressed patients in either case, oncologic patients, partially treated with antibiotics before ED presentation, sustained systemic organ failure/injury, or sterile inflammatory disease.

### DATA COLLECTION AND QUALITY CONTROL

Once patients with suspected infections fulfil the study criterion, single blood (either venous or arterial) sampling was performed for each interest biomarker (high-sensitive CRP, sPLA2-IIA, N%, PCT, and Lactate) according to routine ED standard protocol before medical treatment. For each patient, a total of 5mL of whole blood was collected with 2.5mL dispensed into an EDTA tube for a full blood count. Another 3mL whole blood was withdrawn to the serum tube for high-sensitive CRP, sPLA2-IIA, and PCT. The samples were centrifuged immediately and stored at −85 ° C until the moment of analysis. The serum lactate level was detected from the site lab arterial blood gas results, whereas N% was traced from full blood count results. Relevant culture and serology tests were ordered as determined by the treating physician based on a case-to-case basis.

### METHOD OF DETERMINATION OF BIOMARKERS

The high-sensitive latex immunoassay MULTIGENT CRP Vario with a measurement range of 0.10-160.00mg/L was used to measure high-sensitive CRP levels. For sPLA2-IIA serum activity, the samples were tested in triplicates on sPLA2-IIA (human type IIA) Enzyme Immunometric Assay Kit (Cayman Chemical, USA) as per the manufacturer’s instructions. N% was determined by flow cytometry analysis on an SYSMEX XN-3000 Analyzer. In vitro quantitative determination of PCT value was done with Elecys BRAHMS PCT, which was utilised as an immunoassay. Lactate levels were measured with ABL 800 BASIC analyser.

### STATISTICAL ANALYSIS

All statistical analysis was accomplished using SPSS™ software, version 24. The recruited patients’ demographic data and causative microorganisms were summarised as frequency (%), mean and standard deviation. Parametric variables with normal distribution were presented as mean ± standard deviation. Otherwise, the median and interquartile values were reported instead. Subsequently, non-parametric variables were tested with the Mann-Whitney U test for two-group comparisons and Kruskal-Wallis tests for multi-group comparisons. A 2-sided *P* value of <0.05 was used in simple comparisons to indicate statistical significance. A 2-sided *P* value of <0.01 was adjusted for multiple comparisons to indicate the statistical significance based on Bonferroni’s correction.^30^ Pearson chi-square, χ^2^ test was used to compare the association of categorical variables. Area under receiver operating characteristic curve (AUROC) was used to assess each biomarker’s performance in discriminating between BI and non-BI. The cut-off point of each biomarker was then determined and used as the reference. The Medcal™ online calculator was used to determine Sn, Sp, positive predictive values (PPV), negative predictive values (NPV), and accuracy with a 95% confidence interval (95%CI) of each biomarker. The accuracy of each parameter was verified by Cohen’s kappa (κ) agreement test. An online sample calculator, “easyROC”, was used to calculate the sample size using PCT as a standard biomarker based on past literature. With a type 1 error of 0.05, a power of 0.8, an AUROC PCT of 0.93 from the Luzzani et al. (2003), and an AUROC sPLA2-IIA of 0.93 from Tan et al. (2016) at the lower difference of 0.01, we calculated a total sample size of 112.^12,31^

## CONCLUSION

Taking it all together, sPLA2-IIA is comparable to high-sensitive CRP but better than N%, PCT, and lactate in identifying BI at a fast-paced ED setting in LMIC. Combinations of biomarkers can yield better diagnostic performance on bacterial infection management in LMICs. Further studies should be carried out to explore the potency of sPLA2-IIA, particularly validating its implicit roles as a potential AMS biomarker. This would expedite the physicians’ decision-making for proper antimicrobial administration and mitigate the rising threat of antimicrobial resistance globally.

## Data Availability

All the relevant data was already reported in the manuscript. The complete data that supported the findings of this study are available upon request from the corresponding author (T.L.T.) in view of the patient's personal data privacy and its confidentiality.

## DATA AVAILABILITY

The data that support the findings of this study are available from Toh Leong Tan but restrictions apply to the availability of these data, which were used under license for the current study, and so are not publicly available. Data are however available from the authors upon reasonable request and with permission of Toh Leong Tan.

## ACKNOWLEDGEMENTS

We would also like to thank Universiti Kebangsaan Malaysia and the Ministry of Education, Malaysia, for supporting this research. We thank all the Doctors & EMS personnel, the ED, and all the medical laboratory technicians from the Department of Pathology Universiti Kebangsaan Malaysia Medical Centre.

## AUTHOR INFORMATION

### Affiliations

Department of Emergency Medicine, Faculty of Medicine, Universiti Kebangsaan Malaysia Medical Centre, 56000 Cheras, Kuala Lumpur, Malaysia

Toh Leong Tan, Nurul Saadah Ahmad, Kai Shen Ooi, Swee Thian Tan

Department of Pathology, Faculty of Medicine, Universiti Kebangsaan Malaysia Medical Centre, 56000 Cheras, Kuala Lumpur, Malaysia

Dian Nasriana Nasuruddin, Azlin Ithnin

Department of Biochemistry, Faculty of Medicine, Universiti Kebangsaan Malaysia Medical Centre, 56000 Cheras, Kuala Lumpur, Malaysia

Khaizurin Tajul Arifin

Centre of Advanced Materials and Renewable Resources, Faculty of Science & Technology, Universiti Kebangsaan Malaysia, 43600 Bangi, Selangor, Malaysia

Yook Heng Lee, Nurul Izzaty Hassan

Faculty of Engineering and Built Environment, Universiti Kebangsaan Malaysia, 43600 Bangi, Selangor, Malaysia

Kok Beng Gan

UKM Medical Molecular Biology Institute (UMBI), Universiti Kebangsaan Malaysia Medical Centre, 56000 Cheras, Kuala Lumpur, Malaysia

Hui-min Neoh

Department of Emergency Medicine, Tengku Ampuan Rahimah Hospital, Ministry of Health, 41200 Klang, Selangor, Malaysia

Christabel Wan-li Kang

## Contributions

Conceptualisation, T.L.T., and C.W.K.; Methodology, T.L.T., C.W.K. and N.S.A.; Software, T.L.T., C.W.K., K.S.O., S.T.T., and N.S.A.; Validation, T.L.T., C.W.K., K.S.O. S.T.T., and D.N.N.; Formal Analysis, T.L.T., C.W.K., K.S.O. and S.T.T.; Investigation, T.L.T., C.W.K, K.S.O., S.T.T., N.S.A. D.N.N. A.I., and K.T.A.; Resources, T.L.T., C.W.K., K.S.O., S.T.T. and N.S.A.; Data Curation, T.L.T., C.W.K, K.S.O., S.T.T., N.S.A., D.N.N. and A.I.; Writing — Original Draft Preparation, T.L.T. C.W.K, K.S.O., S.T.T. and H.M.N.; Writing — Review & Editing, T.L.T., C.W.K., K.S.O., S.T.T., N.S.A., D.N.N., A.I., K.T.A., Y.H.L., N.I.H., K.B.G and H.M.N.; Visualisation, T.L.T., K.S.O. and S.T.T.; Supervision, T.L.T.; Project Administration, T.L.T. and N.S.A.; Funding Acquisition, T.L.T, and D.N.N. All authors have read and agreed to the published version of the manuscript.

## ADDITIONAL INFORMATION

### Publisher’s note

Springer Nature remains neutral with regard to jurisdictional claims in published maps and institutional affiliations.

## REFERENCES

1 Wang, H. et al.. Global, regional, and national life expectancy, all-cause mortality, and cause-specific mortality for 249 causes of death, 1980–2015: a systematic analysis for the Global Burden of Disease Study 2015. The lancet 388, 1459–1544, doi:https://doi.org/10.1016/S0140-6736(16)31012-1 (2016).

2 Dadonaite, B. & Roser, M. Pneumonia. Published online at OurWorldInData.org. Retrieved from: https://ourworldindata.org/pneumonia (2018)

3 Aston, S. J. Pneumonia in the developing world: Characteristic features and approach to management. Respirology 22, 1276–1287, doi:https://doi.org/10.1111/resp.13112 (2017).

4 Zar, H. J., Madhi, S. A., Aston, S. J. & Gordon, S. B. Pneumonia in low and middle income countries: progress and challenges. Thorax 68, 1052–1056, doi:10.1136/thoraxjnl-2013-204247 (2013).

5 Department of Statistic Malaysia. Statistics on Causes of Death, Malaysia, 2020 https://www.dosm.gov.my/v1/index.php?r=column/pdfPrev&id=QTU5T0dKQ1g4MHYxd3ZpMzhEMzdRdz09 (2020).

6 Tsao, Y.-T. et al.. Differential Markers of Bacterial and Viral Infections in Children for Point-of-Care Testing. Trends in Molecular Medicine 26, 1118–1132, doi:10.1016/j.molmed.2020.09.004 (2020).

7 Saleh, M. A. A., van de Garde, E. M. W. & van Hasselt, J. G. C. Host-response biomarkers for the diagnosis of bacterial respiratory tract infections. Clinical Chemistry and Laboratory Medicine (CCLM) 57, 442–451, doi:https://doi.org/10.1515/cclm-2018-0682 (2019).

8 Hausfater, P. Biomarkers and infection in the emergency unit. Médecine et Maladies Infectieuses 44, 139–145, doi:https://doi.org/10.1016/j.medmal.2014.01.002 (2014).

9 Kapasi, A. J., Dittrich, S., González, I. J. & Rodwell, T. C. Host biomarkers for distinguishing bacterial from non-bacterial causes of acute febrile illness: a comprehensive review. PLoS One 11, e0160278 (2016).

10 Ljungström, L. et al.. Diagnostic accuracy of procalcitonin, neutrophil-lymphocyte count ratio, C-reactive protein, and lactate in patients with suspected bacterial sepsis. PloS one 12, e0181704, doi:https://doi.org/10.1371/journal.pone.0181704 (2017).

11 Yusa, T., Tateda, K., Ohara, A. & Miyazaki, S. New possible biomarkers for diagnosis of infections and diagnostic distinction between bacterial and viral infections in children. Journal of Infection and Chemotherapy 23, 96–100, doi:10.1016/j.jiac.2016.11.002 (2017).

12 Tan, T. L. et al.. CD64 and group II secretory phospholipase A2 (sPLA2-IIA) as biomarkers for distinguishing adult sepsis and bacterial infections in the emergency department. PloS one 11, e0152065, doi:https://doi.org/10.1371/journal.pone.0152065 (2016).

13 Tan, T. L. & Goh, Y. Y. The role of group IIA secretory phospholipase A2 (sPLA2-IIA) as a biomarker for the diagnosis of sepsis and bacterial infection in adults—A systematic review. PloS one 12, e0180554, doi:https://doi.org/10.1371/journal.pone.0180554 (2017).

14 Rintala, E. M. & Nevalainen, T. J. Group II phospholipase A2 in sera of febrile patients with microbiologically or clinically documented infections. Clinical infectious diseases 17, 864–870, doi:https://doi.org/10.1093/clinids/17.5.864 (1993).

15 Nik Mansor, N. N. et al. An Amperometric Biosensor for the Determination of Bacterial Sepsis Biomarker, Secretory Phospholipase Group 2-IIA Using a Tri-Enzyme System. Sensors 18, 686 (2018).

16 Simon, L., Gauvin, F., Amre, D. K., Saint-Louis, P. & Lacroix, J. Serum procalcitonin and C-reactive protein levels as markers of bacterial infection: a systematic review and meta-analysis. Clinical infectious diseases 39, 206–217, doi:https://doi.org/10.1086/421997 (2004).

17 Bador, K., Intan, S., Hussin, S. & Gafor, A. H. A. Serum procalcitonin has negative predictive value for bacterial infection in active systemic lupus erythematosus. Lupus 21, 1172–1177, doi:10.1177/0961203312450085 (2012).

18 Massaro, K. S., Costa, S. F., Leone, C. & Chamone, D. A. Procalcitonin (PCT) and C-reactive protein (CRP) as severe systemic infection markers in febrile neutropenic adults. BMC infectious diseases 7, 137, doi:https://doi.org/10.1186/1471-2334-7-137 (2007).

19 Gao, L. et al.. Early diagnosis of bacterial infection in patients with septicopyemia by laboratory analysis of PCT, CRP and IL-6. Experimental and therapeutic medicine 13, 3479–3483, doi:10.3892/etm.2017.4417 (2017).

20 Cui, N., Zhang, H., Chen, Z. & Yu, Z. Prognostic significance of PCT and CRP evaluation for adult ICU patients with sepsis and septic shock: retrospective analysis of 59 cases. Journal of International Medical Research 47, 1573–1579, doi:10.1177/0300060518822404 (2019).

21 Escadafal, C. et al.. New Biomarkers and Diagnostic Tools for the Management of Fever in Low- and Middle-Income Countries: An Overview of the Challenges. Diagnostics 7, 44 (2017).

22 Lubell, Y. et al.. Modelling the Impact and Cost-Effectiveness of Biomarker Tests as Compared with Pathogen-Specific Diagnostics in the Management of Undifferentiated Fever in Remote Tropical Settings. PLoS One 11, e0152420, doi:10.1371/journal.pone.0152420 (2016).

23 Lubell, Y. et al.. Performance of C-reactive protein and procalcitonin to distinguish viral from bacterial and malarial causes of fever in Southeast Asia. BMC infectious diseases 15, 511, doi:10.1186/s12879-015-1272-6 (2015).

24 Escadafal, C., Incardona, S., Fernandez-Carballo, B. L. & Dittrich, S. The good and the bad: using C reactive protein to distinguish bacterial from non-bacterial infection among febrile patients in low-resource settings. BMJ Global Health 5, e002396, doi:10.1136/bmjgh-2020-002396 (2020).

25 Zakariah, N. A. et al.. Is Procalcitonin more superior to hs-CRP in the diagnosis of infection in diabetic foot ulcer? The Malaysian journal of pathology 42, 77–84 (2020).

26 Schuetz, P. et al.. Procalcitonin (PCT)-guided antibiotic stewardship: an international experts consensus on optimized clinical use. Clinical Chemistry and Laboratory Medicine (CCLM) 57, 1308–1318, doi:doi:10.1515/cclm-2018-1181 (2019).

27 Lee, C.-C. et al.. Procalcitonin (PCT)-guided antibiotic stewardship in Asia-Pacific countries: adaptation based on an expert consensus meeting. Clinical Chemistry and Laboratory Medicine (CCLM) 58, 1983–1991, doi:doi:10.1515/cclm-2019-1122 (2020).

28 Wasserman, M., Levinstein, M., Keller, E., Lee, S. & Yoshikawa, T. T. Utility of fever, white blood cells, and differential count in predicting bacterial infections in the elderly. Journal of the American Geriatrics Society 37, 537–543, doi:https://doi.org/10.1111/j.1532-5415.1989.tb05686.x (1989).

29 Horan, T. C., Andrus, M. & Dudeck, M. A. CDC/NHSN surveillance definition of health care–associated infection and criteria for specific types of infections in the acute care setting. American journal of infection control 36, 309–332, doi:10.1016/j.ajic.2008.03.002 (2008).

30 Bland, J. M. & Altman, D. G. Multiple significance tests: the Bonferroni method. Bmj 310, 170, doi:10.1136/bmj.310.6973.170 (1995).

31 Luzzani, A. et al.. Comparison of procalcitonin and C-reactive protein as markers of sepsis. Critical care medicine 31, 1737–1741, doi:10.1097/01.CCM.0000063440.19188.ED (2003).

